# Molecular Imaging of Androgen Receptor Expression and Glycolysis as Biomarkers for Clinical Outcome for Metastatic Castration–Resistant Prostate Cancer

**DOI:** 10.1101/2025.06.06.25329162

**Authors:** Samir Zaidi, Jessica Flynn, Josef J. Fox, Andreas G. Wibmer, Glenn Heller, Howard I. Scher, Steven M. Larson, Michael J. Morris

**Author notes:** **Conflict of interest:** None. **Funding:** Prostate Cancer Foundation (SZ and MJM). <u>Corresponding Author</u>: Samir Zaidi, MD, PhD Assistant Attending Physician Genitourinary Oncology Service, Department of Medicine Memorial Sloan Kettering Cancer Center 1275 York Avenue New York, NY 10065 (now at Yale Cancer Center).

## Abstract

**Purpose:** Patients with metastatic castration–resistant prostate cancer (mCRPC) with tumors that display disease heterogeneity, including features of androgen receptor (AR)–independence and/or lineage plasticity, have particularly poor outcomes. Non–invasive methods to prognosticate on such poor–risk features continue to pose a major challenge. We previously showed that imaging could be used to quantitatively identify patients by virtue of AR–expression and glycolysis, and that those with AR–negative and glycolytic–positive disease were associated with a poorer prognosis. Here, we extend our analysis by creating a model integrating imaging parameters with clinical determinants to comprehensively prognosticate progression free (PFS) and overall survival (OS).

**Patients and Methods:** 124 CRPC patients underwent dual FDG/FDHT PET scans in a clinical trial spanning 7–years. For each patient, clinical markers and five index lesions showing abnormal tracer uptake and anatomical tumor spread were analyzed. Univariate analysis was performed on PET and clinical variables for OS and PFS. Cox proportional–hazard models were developed for OS and PFS with an analysis of covariance determining association between FDG/FDHT and OS/PFS, after adjustment for serum markers. Concordance probability estimates (CPE) quantified the discriminatory power of our model.

**Results:** FDG SUV_maxavg_ uptake was associated with OS (HR=1.55, *P*=0.027) with the 4^th^ quartile of FDG SUV_maxavg_ (≥7.8) showing a 14–month reduction in OS relative to the 1^st^ quartile (≤4.22). Moreover, through multivariate modeling, the incorporation of FDG PET with clinical markers yielded a model with moderate discriminatory power (CPE=0.74). FDHT SUV_maxavg_ uptake was however not associated with OS (HR=1.14, *P*=0.5). Furthermore, FDHT SUV_max_ as a covariate lacked robust association with PFS (HR=0.78, *P*=0.08).

**Conclusions:** In this prognostic model that integrates clinical and molecular imaging data, FDG PET is an important element of identifying poor–risk patients by OS. FDHT, however, does not prognosticate for OS, and further lacks robust association with PFS, likely due to the complexities of imaging AR in mCRPC patients.

## INTRODUCTION

Despite the efficacy of second–generation androgen receptor signaling inhibitors (ARSI’s) (*1–4*) used for both castration–naïve and castration–resistant disease, most patients with metastatic prostate cancer eventually develop treatment resistance and die of disease. In one resistance mechanism, which is of growing importance, cancer cells adopt an alternative AR– independent gene program, often referred to as lineage plasticity (*5–7*). Tumors undergoing lineage plasticity are enriched for *TP53* and *RB1* alterations and comprise a range of AR–negative adenocarcinomas and high–grade mixed neuroendocrine cancers (*8–10*). An increasing incidence of these cancers with the use of androgen receptor signaling inhibitors (ARSIs), along with the growing recognition of significant disease heterogeneity in late stage mCRPC, has prompted trials focused on drugs targeting lineage plasticity, particularly as these patients have the worst clinical outcomes (*11*). However, given that there is no consensus for identifying or classifying these tumor types, patient selection for clinical studies and the choice of therapeutic regimens remains an ongoing dilemma. There is thus a major unmet need to identify robust prognostic tools in the context of such tumor heterogeneity to inform treatment decisions.

Towards this goal, molecular imaging based on positron emission tomography (PET) has provided the sensitivity and precision for prostate cancer detection (*12–18*). [^18^F]– fluorodeoxyglucose (FDG) imaging, a measure of glycolysis, has shown promise as a prognostic and predictive biomarker in mCRPC in well controlled clinical studies (*13, 14, 19, 20*). More recently, prostate specific membrane antigen (PSMA) PET imaging has been approved as a companion biomarker for treatment with Lutetium-177-PMSA-617 in mCRPC (*21*). A recent biomarker analysis demonstrated that both PSMA and FDG PET imaging are predictive biomarkers in patients given Lutetium-177-PMSA-617 (*22*). Furthermore, we and others have shown that in FDG–avid disease in the absence of prostate–specific imaging using [^68^Ga]– prostate–specific membrane antigen (PSMA) or [^18^F]–16β–fluoro-5α-dihydrotestosterone (FDHT) conveys a poor prognosis (*16, 23*). Our earlier study documenting individual patient–level lesion heterogeneity using both FDG and FDHT has further suggested the additional importance of measuring AR in a quantitative manner (*12, 15, 16*). Here, our study seeks to further understand whether baseline FDG and FDHT PET imaging parameters in addition to histologic and serum biomarkers prognosticate overall survival (OS) and progression–free survival (PFS) in men with CRPC in a clinical trial spanning a 7–year period.

## PATIENTS AND METHODS

### Patient Selection

Patients with progressive mCRPC were enrolled to undergo FDG and FDHT PET/CT imaging as part of a prospective clinical trial (NCT00588185) with patients accrued between 2007 and 2013. Clinical protocols were reviewed and approved by Memorial Sloan Kettering Cancer Center (MSKCC) Institutional Review Board. Participants provided written informed consent, and there was no financial compensation. Eligible patients were required to have progressive, histologically confirmed CRPC and visible lesions on standard computed tomography (CT), magnetic resonance imaging (MRI), or bone scintigraphy consistent with metastatic disease. Progressive disease was defined as a rising PSA (25% increase over 3 weekly or 2 bimonthly observations), new lesions on bone scintigraphy, or an increase in soft tissue disease (or new lesions) on trans–axial imaging. Between 2007 to 2013, 173 patients consented to NCT00588185, of which 49 patients were excluded for either not having both FDHT and FDG PET scans or starting treatment >45 days after baseline scans. This yielded a total of 124 patients analyzed in this study.

The baseline evaluation included complete blood counts, PSA, testosterone, dihydrotestosterone, blood urea nitrogen, creatinine, hepatic transaminases, bilirubin, alkaline phosphatase, and albumin. Exclusion criteria included: bilirubin >1.5 x upper limit of normal (ULN); aspartate transaminase/alanine transaminase (AST/ALT) >2.5 x ULN; albumin <2 g/dl; gamma–glutamyl transpeptidase (GGT) and alkaline phosphatase >2.5 x ULN; creatinine >1.5 x ULN; and/or a creatinine clearance <60 mL/min. In addition to FDG and FDHT PET/CT, patients also underwent bone scintigraphy, CT or MRI of the abdomen and pelvis, and at least a chest radiograph within 6 weeks before or 2 weeks after study entry. Treatment after baseline scans included next–generation hormonal therapy (66 patients on abiraterone, enzalutamide or apalutamide), chemotherapy (4 patients on paclitaxel, carboplatin or docetaxel), immunotherapy (1 patient given CAR–T cells targeted to PSMA), investigational trials (with some patients on hormonal–based therapy), radiation–based treatments (4 patients), and other therapies (7 patients received ketoconazole, dutasteride or estradiol).

### Imaging Parameters

Prior to 2007, images were obtained from the skull to mid thighs on a dedicated PET GE Advance Scanner, and on GE Discovery LS and Siemens Biograph PET/CT scanners. From 2007 onwards, all PET/CT scans were acquired on a GE Discovery STE PET/CT scanner. PET reconstruction and cross calibration between PET scanners to assure quantitative equivalence and preparation of FDG and FDHT was performed as previously described (*16*). Patients underwent FDG and FDHT imaging on separate days. With the exception of water, patients fasted for 6 hours before injection with ∼10 mCi (370 MBq) FDG. FDHT was prepared at MSKCC as previously described (*16*), with ∼333 Mbq (9 mCi) being injected without dietary preparation. Images were obtained ∼60 and ∼40 minutes after FDG or FDHT injections, respectively.

### Image Analysis and Quantitation

PET scans were read by reference radiologists who were unaware of bone scintigraphy findings. Five index lesions indicative of abnormal tracer uptake and representative of the anatomical spread of disease were selected for semiquantitative analysis. Special consideration was given to select lesions from diverse anatomic areas in order to represent true disease spread accurately. SUV_max_ represented the lesion with the highest standardized uptake value of the five index lesions. SUV_maxavg_ was computed as the average of these 5 index lesions (*12–16*). SUVs were obtained using the following formula:

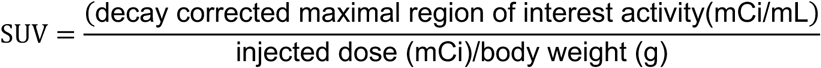

Index lesions sites were recorded and categorized into 6 sites of disease: bone, lung, liver, lymph node, prostate/prostate mass, or other. Patients underwent baseline FDG and FDHT PET imaging during disease progression and prior to starting a new therapeutic regimen.

### Statistical Analysis

OS was defined from date of first therapeutic treatment following the baseline PET scans to last follow–up or death. PFS was defined from first therapeutic treatment following the baseline PET scans until the first PSA rise (25% increase over 3 weekly or 2 bimonthly observations) or death. Univariate analysis was performed on all PET variables and lab markers for both OS and PFS endpoints using Cox regression models. A nonparametric kernel smoothed relation between the median OS or PFS and FDG, and FDHT SUV_max_ or SUV_maxavg_ were generated using the *‘sm’* package in R. The FDG and FDHT SUV_max_ or SUV_maxavg_ were grouped into quartiles and Kaplan Meier estimates of OS and PFS by group were computed. Cox proportional hazards models were developed for OS and PFS using a backward variable selection procedure with a retaining criterion of *P*<0.05. The risk factors considered in the models were: log PSA, albumin, hemoglobin, log alkaline phosphatase, log lactate dehydrogenase, Gleason score, total lesion number, and FDG and FDHT log SUV_max_ and log SUV_maxavg_. The log transformation of some of these factors was due to their right skewness. An analysis of covariance, based on the proportional hazards model, was used to determine the association between FDG/FDHT measures and OS/PFS after adjusting for relevant risk factors. Concordance probability estimates (CPEs) were determined for the Cox models to quantify the discriminatory power of a specific model.

## RESULTS

A total of 173 CRPC patients who underwent dual (FDG and FDHT PET) scans were selected during this enrollment period. With 49 excluded (refer to Methods), we analyzed a total of 124 mCRPC patients with dual PET imaging. Baseline patient characteristics and treatment are shown in **Table 1**. Median age was 67 years (range: 44 to 85). Histopathologic review revealed that 63% had ≥ Gleason 8 tumors with a median PSA of 45 (range: 1 to 1477 ng/mL). Patients were largely in good performance status, with only 4% showing a Karnofsky Performance Status of <80. All patients had received prior hormonal therapies, with 49% and 13% also having received chemotherapy and immunotherapy, respectively. All met the protocol requirement to start a new treatment within 45 days of imaging, median 1.1 weeks (range: 0 to 6 weeks), which included AR–directed therapies (56%, 69 patients: 66 in hormonal/AR–directed and 3 in investigational groups) (**Table 1**). The median follow–up was 21.2 months (range: 1.3 to 114.3 months).

**Table 1.** Patient Demographics.

| **Characteristic** | **N = 124** |
| --- | --- |
| Age, years | 67 (44, 85) |
| Gleason score |  |
| <8 | 45 (37%) |
| >=8 | 77 (63%) |
| Unknown | 2 |
| KPS |  |
| 70 | 5 (4.1%) |
| 80 | 52 (42%) |
| 90 | 66 (54%) |
| Unknown | 1 |
| PSA, ng/mL | 45 (1, 1477) |
| Hemoglobin, g/dL | 12.4 (8.9, 15.6) |
| LDH, U/L | 209 (109, 1404) |
| Unknown | 4 |
| Alkaline Phosphatase, U/L | 95 (36, 2148) |
| Albumin, g/dL | 4.3 (3.2, 5) |
| Prior Chemotherapy | 61 (49%) |
| Prior Immunotherapy | 16 (13%) |
| No. Prior Hormonal or AR Directed Treatments |  |
| 1 | 23 (19%) |
| 2 | 45 (37%) |
| 3 | 31 (25%) |
| >3 | 23 (19%) |
| Unknown | 2 |
| Total Lesion Number | 16 (1, 61) |
| Treatment Type |  |
| Hormonal/AR-directed | 66 (53%) |
| Chemotherapy | 4 (3.2%) |
| Immunotherapy | 1 (0.8%) |
| Investigational* | 42 (34%) |
| Radiation | 4 (3.2%) |
| None | 4 (3.2%) |
| Other | 3 (2.4%) |

Statistics presented: median (minimum, maximum); n (%)
\*3 investigational treatments included hormonal or AR directed treatment

To evaluate both FDG and FDHT lesions, we determined the average and highest standardized uptake values (SUV_maxavg_ and SUV_max_) of five index lesions for each patient (refer to **Methods** for description of index lesion selection) (*14*). The median OS within the cohort was 21.2 months with a 5–year survival of 11%, consistent with prior studies (**Fig. 1A**) (*1, 4*). Serologically, and in addition to PSA (HR,1.25; *P*<0.001, log scale), hemoglobin (HR, 0.83; *P*=0.006), lactate dehydrogenase (HR, 9.28; *P*<0.001, log scale), alkaline phosphatase (HR, 1.9; *P*<0.001, log scale), and albumin (HR, 0.44, *P*=0.02) were prognostic of OS using the Wald test from the proportional hazards model (**Table 2**). Total lesion number was also associated with poor survival (HR, 1.04, *P*≤0.001) (**Table 2**). For PET FDG scans, high SUV_maxavg_ and SUV_max_ showed evidence of a poorer overall survival (HR=1.43, *P*=0.05 and HR, 1.31, *P*=0.07, respectively, log scale). In contrast, for FDHT scans, we could not find a relationship between SUV_maxavg_ or SUV_max_ and OS (HR 1.07; *P*=0.5; and HR 1.14, *P*=0.5, respectively) (**Table 2**).

**Figure 1.**
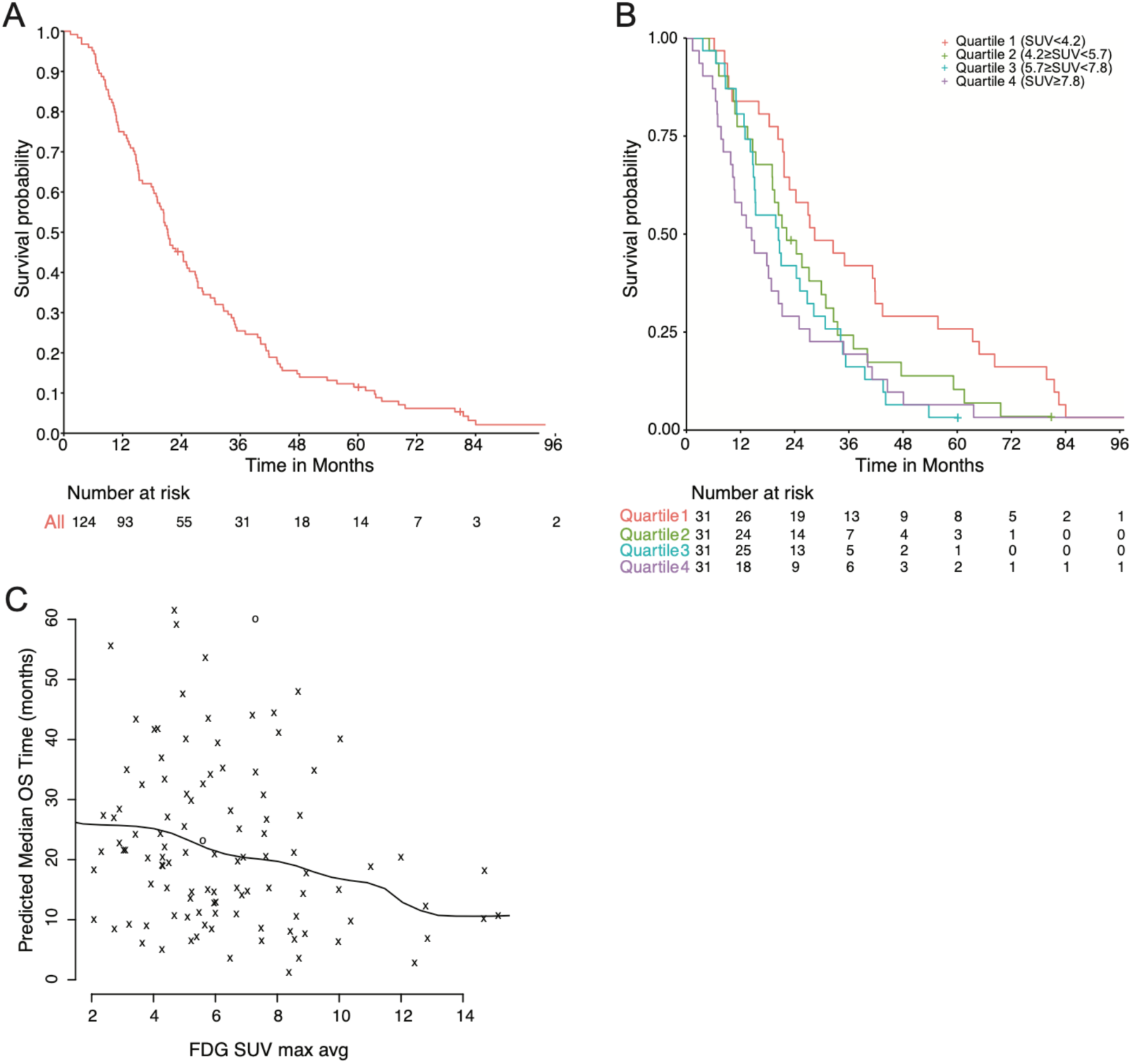
Association of FDG PET SUV values with overall survival (OS) in CRPC patients. **(A)** OS is shown for 124 CRPC analyzed patients, who underwent baseline FDG PET and FDHT imaging. Median OS is 21.2 months. **(B)** FDG SUV_maxavg_ was separated in quartiles for OS using Cox regression models with median OS calculated for each SUV quartile (Q1, 28 months; Q2, 22 months; Q3, 20 months; and Q4, 14 months). **(C)** Modeling of FDG SUV_maxavg_ as a continuous variable. Values are plotted on a scatter plot with median OS and with smoothed curves.

**Table 2.**
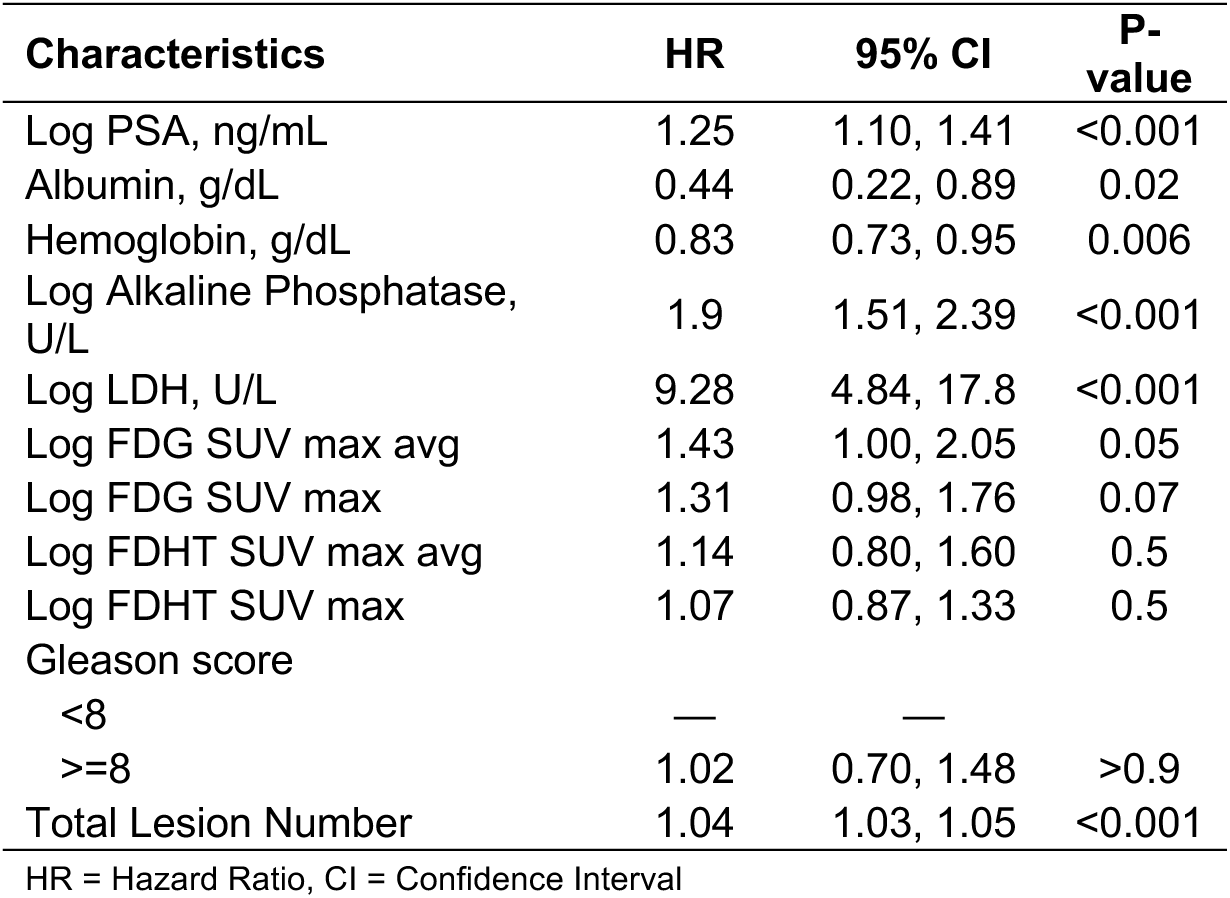
Univariate Analysis for Survival.

| Characteristics | HR | 95% CI | P-value |
| --- | --- | --- | --- |
| Log PSA, ng/mL | 1.25 | 1.10, 1.41 | <0.001 |
| Albumin, g/dL | 0.44 | 0.22, 0.89 | 0.02 |
| Hemoglobin, g/dL | 0.83 | 0.73, 0.95 | 0.006 |
| Log Alkaline Phosphatase, U/L | 1.9 | 1.51, 2.39 | <0.001 |
| Log LDH, U/L | 9.28 | 4.84, 17.8 | <0.001 |
| Log FDG SUV max avg | 1.43 | 1.00, 2.05 | 0.05 |
| Log FDG SUV max | 1.31 | 0.98, 1.76 | 0.07 |
| Log FDHT SUV max avg | 1.14 | 0.80, 1.60 | 0.5 |
| Log FDHT SUV max | 1.07 | 0.87, 1.33 | 0.5 |
| Gleason score |  |  |  |
| <8 | — | — |  |
| >=8 | 1.02 | 0.70, 1.48 | >0.9 |
| Total Lesion Number | 1.04 | 1.03, 1.05 | <0.001 |

HR = Hazard Ratio, CI = Confidence Interval

To further depict the relationship between FDG SUV_maxavg_ values and OS, we grouped FDG SUV_maxavg_ into quartiles and calculated the median overall survival within each quartile (**Fig. 1B**). Consistent with our data, the fourth quartile (cutoff: FDG SUV_maxavg_≥SUV=7.8, mOS=14 months) showed a 14–month reduction in median OS compared to the first quartile (cutoff: FDG SUV_maxavg_<SUV**=**4.22, mOS=28 months). Similarly, we modeled FDG SUV_maxavg_ as a continuous variable, which revealed similar evidence of higher FDG SUV_maxavg_ values being associated with a lower survival (**Fig. 1C).**

Building upon our prior work, we implemented an analysis of covariance using the proportional hazards model to evaluate whether FDG SUV_maxavg_ after adjustment with validated clinical biomarkers was associated with OS (refer to **Methods**). This analysis demonstrated that FDG SUV_maxavg_ was prognostic of OS (HR, 1.55; *P*=0.027). In addition, the Cox model indicated that traditional biomarkers, including serum albumin (HR, 0.39, *P*=0.012) and serum LDH (HR, 6; *P*<0.001) were associated with OS. Furthermore, the CPE value of this model was 0.74, suggestive of moderate discriminatory power (**Table 3**). An example of a CRPC patient identified by our multivariate model with poor prognostic features is shown in **Fig. 2**. This patient showed a FDG SUV_maxavg_ value in the fourth quartile (refer to **Fig. 1B**) with all metabolic lesions demonstrating “concordant” AR and glycolysis activity (**Fig. 2**).

**Figure 2.**
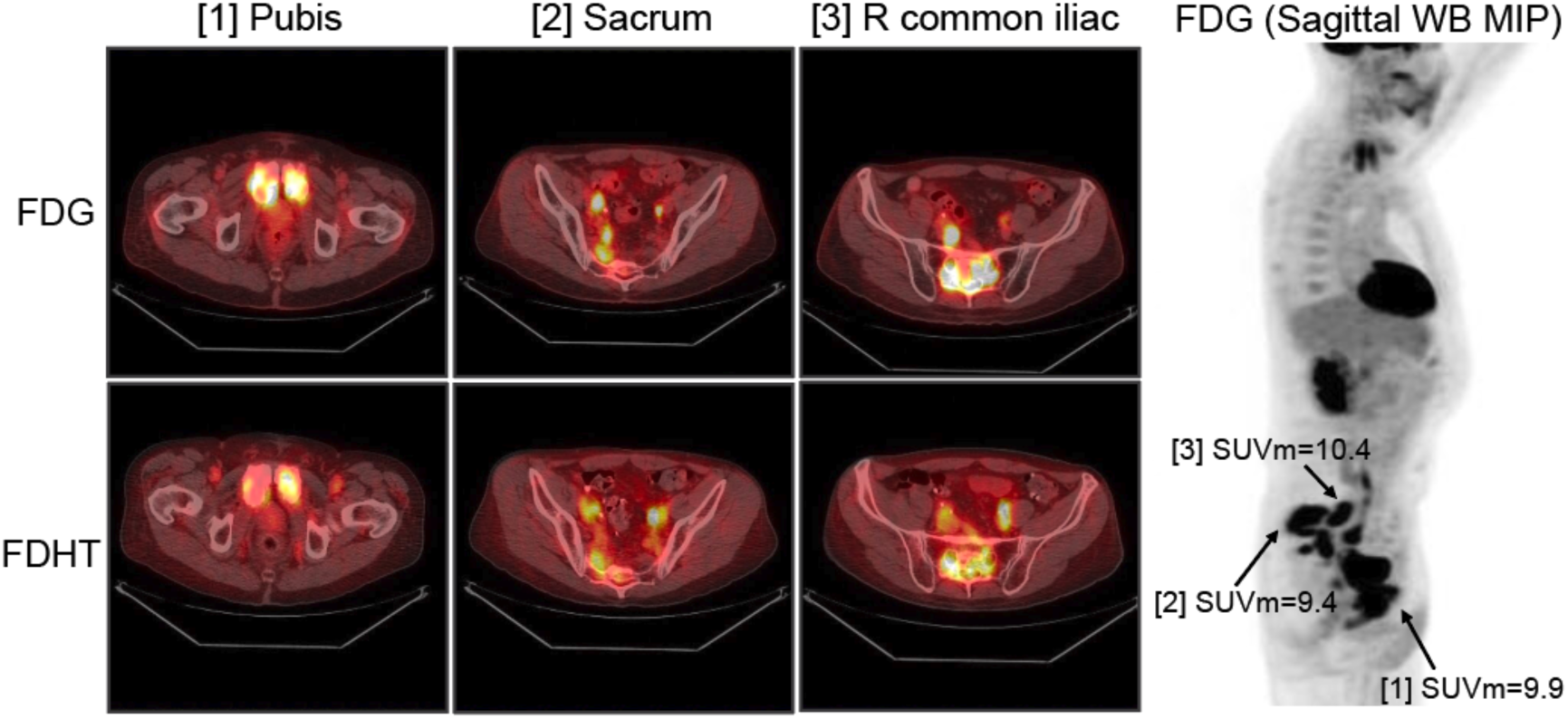
FDG and FDHT Imaging in Patient with Poor Prognostic Features. For both FDG and FDHT, imaging commenced using Discovery 690 PET/CT. Abnormalities are confined to the pelvis with metastatic disease noted in lymph nodes and pelvic bones. Fusion images are shown, which combine PET imaging and CT images obtained during the PET/CT examination, at 3 cross-sectional levels of active disease within pelvis and lower abdomen: 1) at level of pubic synthesis, 2) level of common iliac lymph nodes and 3) level of sacrum. Overall, lesions have roughly comparable SUV uptake of FDG and FDHT, reflecting accelerated glycolysis and AR expression, an imaging phenotype we refer to as “concordant”. Standardized uptake values (SUV) maximum are noted. Of note, there is extensive bone invasion in the sacrum and bilateral pubic rami. Soft tissue involvement is seen in the common iliac and deep pelvic nodes, such as right obturator. Overall, the metabolic tumor volume (MTV) in this patient’s body is estimated at 85.6cc, and his prognosis score is in the 4^th^ quartile (Median SUV_max-avg_>7.8) (median overall survival 14 months, refer to legend of Figure 1B).

**Table 3.** Multivariate Analysis on Survival*.

| Characteristics | HR | 95% CI | P-value |
| --- | --- | --- | --- |
| Albumin | 0.39 | 0.19,0.81 | 0.12 |
| Log LDH | 6 | 3.08,11.7 | <0.001 |
| Log FDG SUV max avg | 1.55 | 1.05, 2.28 | 0.027 |
| Total Lesion Number | 1.03 | 1.02,1.05 | <0.001 |
| **CPE = 0.74** |  |  |  |

HR = Hazard Ratio, CI = Confidence Interval
\*Backward variable selection of factors from univariate analysis (P<0.05)

A rising PSA is the most frequent indicator of treatment resistance to hormonal agents and PFS (*21*). We therefore studied the effects of PET imaging modalities on PFS defined as the date of first PSA rise. Our cohort showed a median PFS of 2.76 months (**Fig. 3**). The proportional hazards model revealed that serum PSA (HR, 1.16; *P*=0.015, log scale), LDH (HR, 2.75; *P*<0.001, log scale), and total lesion number (HR, 1.02; *P*=0.006) were prognostic of PFS. Conversely to our overall survival analyses, FDG SUV_max_ did not show a significant association with PFS (HR = 1.1, *P=*0.5). FDHT SUV_max_ did provide some evidence of association with PFS (HR, 0.77, *P*=0.057, respectively) (**Table 4**). However, by incorporating FDHT along with serum LDH in a proportional hazards model, we found that FDHT SUV_max_ lacked robust association with PFS (FDHT SUV_max_ HR, 0.78, *P*=0.08, CPE 0.61) (**Table 5**).

**Figure 3.**
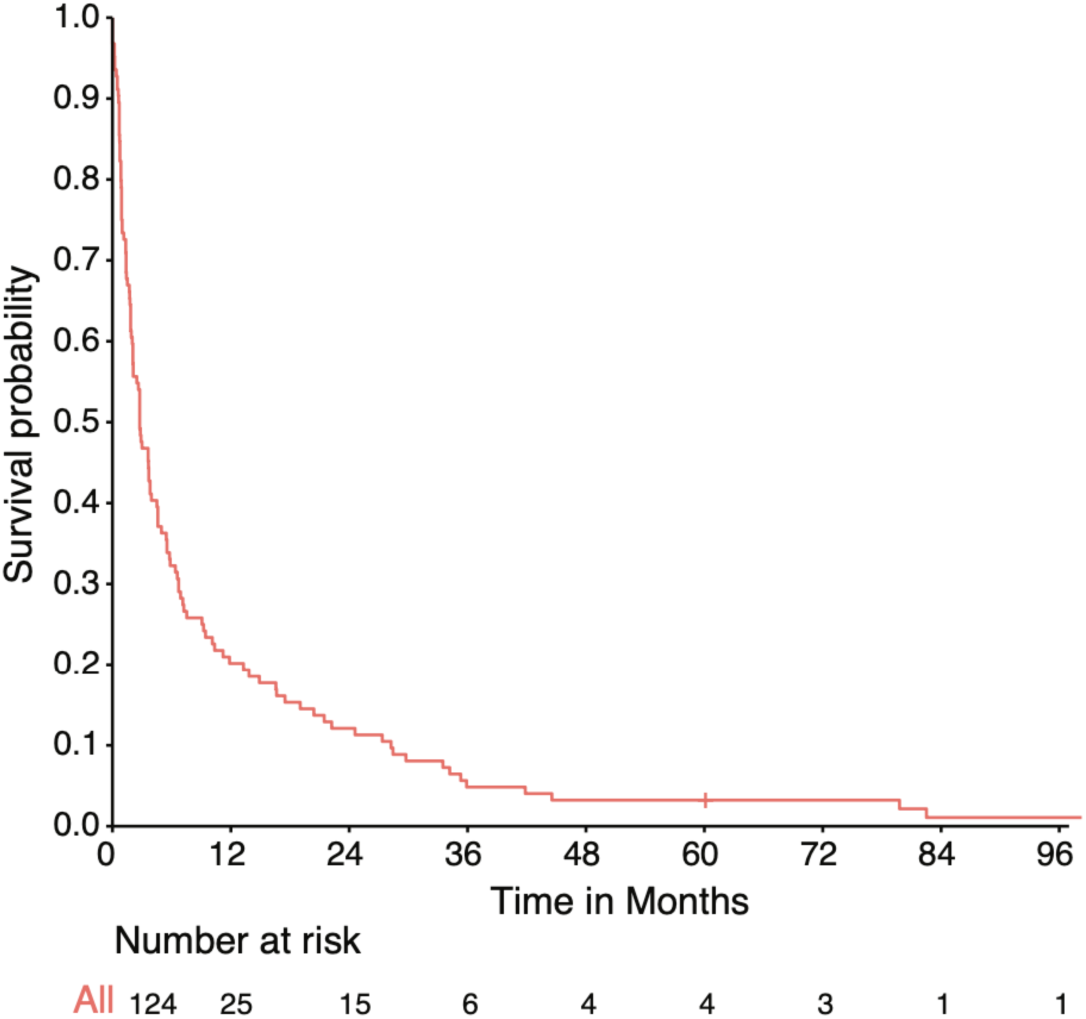
Trend of PET FDHT SUV values with progression free survival (PFS) in CRPC patients. PFS is shown among 124 CRPC analyzed patients, who underwent baseline FDG PET and FDHT imaging. Median OS is 2.76 months.

**Table 4.** Univariate Analysis for PFS.

| Characteristics | HR | 95% CI | P-value |
| --- | --- | --- | --- |
| Log PSA, ng/mL | 1.16 | 1.03, 1.31 | 0.02 |
| Albumin, g/dL | 0.77 | 0.41, 1.46 | 0.4 |
| Hemoglobin, g/dL | 0.93 | 0.82, 1.06 | 0.3 |
| Log Alkaline Phosphatase, U/L | 1.21 | 0.96, 1.53 | 0.1 |
| Log LDH, U/L | 2.75 | 1.7, 4.46 | <0.001 |
| Log FDG SUV max avg | 1.09 | 0.82, 1.46 | 0.5 |
| Log FDG SUV max | 1.1 | 0.84, 1.44 | 0.5 |
| Log FDHT SUV max avg | 0.94 | 0.67, 1.33 | 0.7 |
| Log FDHT SUV max | 0.77 | 0.59, 1.01 | 0.057 |
| Gleason score |  |  |  |
| <8 | — | — |  |
| ≥8 | 0.79 | 0.54, 1.14 | 0.2 |
| Total Lesion Number | 1.02 | 1, 1.03 | 0.006 |

HR = Hazard Ratio, CI = Confidence Interval

**Table 5.**
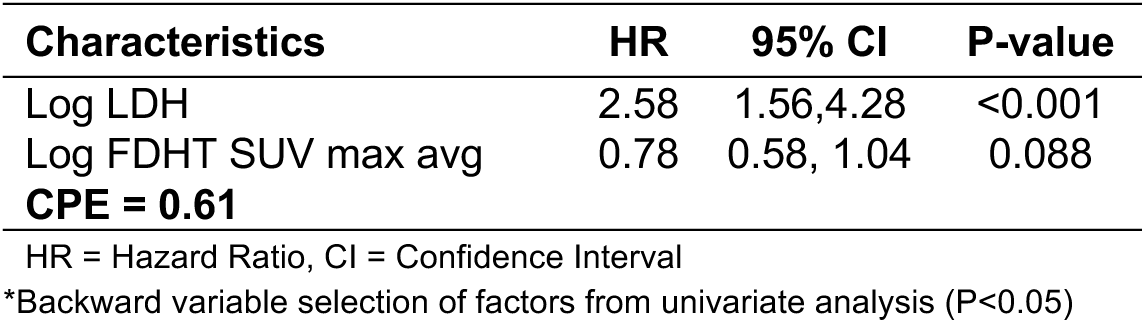
Multivariate Analysis on PFS*.

## DISCUSSION

Most current prognostic models are limited to pathologic, serologic, and clinical biomarkers and do not include direct indicators of disease heterogeneity (*24–26*). FDG PET has been shown to be prognostic for survival. Moreover, our prior data has shown that FDHT together with FDG can yield quantitative measures of tumor heterogeneity and AR–binding at the level of a single lesion with disease heterogeneity being associated with prognosis (*12, 15, 16, 22*). The unmet need, however, is to develop robust multivariate prognostic tools that incorporate both non–invasive imaging modalities together with other validated clinical biomarkers to inform treatment decisions in mCRPC population with high degrees of tumoral heterogeneity. We thus modelled both FDG and FDHT within the context of routine clinical biomarkers to study its role as a prognostic marker for survival in a prospective clinical trial of 124 CRPC patients spanning 7 years.

In our study, FDG SUV uptake showed a strong association with OS, consistent with prior reports (*16, 22*). We also found that FDG SUV uptake is independent of serologic biomarkers in its association with OS. These findings, along with others, demonstrate the role of FDG PET in late stage heterogenous mCRPC disease in prognostication of overall survival and informing treatment decisions. Lastly, multivariate modeling of both FDG PET with clinical markers, namely LDH and albumin, yields a moderate discriminatory index (CPE of 0.74) for prognosticating OS. This will require further confirmation with a validation cohort or an independent prospective clinical trial, but nonetheless highlights the importance of establishing such models that incorporate the entirety of both imaging and clinical biomarkers in prognosticating overall survival.

We also note some evidence of association between FDHT SUV_max_ and PFS suggestive of a subset of these patients potentially having more durable responsiveness to AR directed therapy. This finding will need to be further explored in a cohort with full lesion analysis not limited to the use of SUV_maxavg_ – an average of five index lesions and does not necessarily capture AR– negative lesions that may drive a particularly poor prognosis. We have previously shown that DHT negative, FDG positive lesions (AR_0_Glyc_1_) increase the risk of death by 11% (*16*). This is also consistent with the biologic understanding and clinical evidence that AR loss and lineage plasticity lead to poor clinical outcomes (*9, 27, 28*). We nonetheless note that robust association of FDHT uptake with OS and PFS may be confounded by a variety of factors, including intratumoral endogenous testosterone levels that compete with the tracer for receptor occupancy. In addition, receptor expression levels and constitutively activated receptors without a ligand binding domain can potentially confound the FDHT readout.

Our study confirms the importance of PET FDG as a biomarker for prognosis and illustrates the growing utility and importance of refining multivariate risk models to prognosticate for mCRPC patients with increased risk of death by utilizing both metabolic (FDG) and tumor specific (FDHT and/or PSMA) PET tracers (example shown in **Fig. 2**). We specifically present a potential clinical model that depends heavily on PET imaging for such prognostication that will require independent validation and likely further refinement on a full lesional basis. Such studies will be crucial in informing treatment decisions for mCRPC with significant disease heterogeneity and plasticity.

## CONCLUSIONS

With contemporary treatment paradigms, disease heterogeneity encompassing non– androgen receptor driven prostate cancer is of increasing relevance. To address whether dual imaging with FDG PET and FHT may have prognostic relevance for clinical outcomes, we have analyzed data from a prospective clinical trial spanning 7 years with 124 CRPC patients. By quantitatively measuring index lesions for tracer avidity using both FDG PET and FDHT modalities, we found a strong association of FDG standardized uptake value (SUV) with OS, which when modeled with pathological and serum biomarkers, revealed a utilizable approach for prognosticating OS.

## AUTHOR CONTRIBUTIONS

Conception and design: GH, HIS, SML, MJM. Development, analysis, and methodology: SZ, JF, JJF, AW, GL, HIS, SML, MJM. Writing/review and revision: SZ, JF, GL, SML, MJM. Study supervision: MJM.

## Data Availability

All data is available within the supplement of this paper. If any additional data is needed, please contact corresponding author.

## ACKNOWLEDGMENTS

We are incredibly grateful to the prostate cancer patients who participated in this research. We further appreciate the efforts of the Memorial Sloan Kettering Cancer Center (MSK) Genitourinary faculty and research team. SZ and MJM are supported by the Prostate Cancer Foundation.

## REFERENCES

1. H. I. Scher et al., Increased survival with enzalutamide in prostate cancer after chemotherapy. The New England journal of medicine 367, 1187–1197 (2012).

2. C. J. Ryan et al., Abiraterone in metastatic prostate cancer without previous chemotherapy. The New England journal of medicine 368, 138–148 (2013).

3. J. S. de Bono et al., Abiraterone and increased survival in metastatic prostate cancer. The New England journal of medicine 364, 1995–2005 (2011).

4. T. M. Beer et al., Enzalutamide in metastatic prostate cancer before chemotherapy. The New England journal of medicine 371, 424–433 (2014).

5. S. Y. Ku et al., Rb1 and Trp53 cooperate to suppress prostate cancer lineage plasticity, metastasis, and antiandrogen resistance. Science 355, 78–83 (2017).

6. P. Mu et al., SOX2 promotes lineage plasticity and antiandrogen resistance in TP53- and RB1-deficient prostate cancer. Science 355, 84–88 (2017).

7. H. Beltran et al., Divergent clonal evolution of castration-resistant neuroendocrine prostate cancer. Nat Med 22, 298–305 (2016).

8. M. P. Labrecque et al., Molecular profiling stratifies diverse phenotypes of treatment-refractory metastatic castration-resistant prostate cancer. J Clin Invest 129, 4492–4505 (2019).

9. H. Beltran et al., The Role of Lineage Plasticity in Prostate Cancer Therapy Resistance. Clin Cancer Res 25, 6916–6924 (2019).

10. S. Zaidi et al., Multilineage plasticity in prostate cancer through expansion of stem–like luminal epithelial cells with elevated inflammatory signaling. bioRxiv, 2021.2011.2001.466599 (2021).

11. W. Abida et al., Genomic correlates of clinical outcome in advanced prostate cancer. Proc Natl Acad Sci U S A 116, 11428–11436 (2019).

12. P. B. Zanzonico et al., PET-based radiation dosimetry in man of 18F-fluorodihydrotestosterone, a new radiotracer for imaging prostate cancer. J Nucl Med 45, 1966–1971 (2004).

13. M. J. Morris et al., Fluorinated deoxyglucose positron emission tomography imaging in progressive metastatic prostate cancer. Urology 59, 913–918 (2002).

14. M. J. Morris et al., Fluorodeoxyglucose positron emission tomography as an outcome measure for castrate metastatic prostate cancer treated with antimicrotubule chemotherapy. Clin Cancer Res 11, 3210–3216 (2005).

15. S. M. Larson et al., Tumor localization of 16beta-18F-fluoro-5alpha-dihydrotestosterone versus 18F-FDG in patients with progressive, metastatic prostate cancer. J Nucl Med 45, 366–373 (2004).

16. J. J. Fox et al., Positron Emission Tomography/Computed Tomography-Based Assessments of Androgen Receptor Expression and Glycolytic Activity as a Prognostic Biomarker for Metastatic Castration-Resistant Prostate Cancer. JAMA Oncol 4, 217–224 (2018).

17. M. J. Morris et al., Diagnostic Performance of (18)F-DCFPyL-PET/CT in Men with Biochemically Recurrent Prostate Cancer: Results from the CONDOR Phase III, Multicenter Study. Clin Cancer Res 27, 3674–3682 (2021).

18. K. J. Pienta et al., A Phase 2/3 Prospective Multicenter Study of the Diagnostic Accuracy of Prostate Specific Membrane Antigen PET/CT with (18)F-DCFPyL in Prostate Cancer Patients (OSPREY). J Urol 206, 52–61 (2021).

19. J. P. Buteau et al., PSMA PET and FDG PET as predictors of response and prognosis in a randomized phase 2 trial of 177Lu-PSMA-617 (LuPSMA) versus cabazitaxel in metastatic, castration-resistant prostate cancer (mCRPC) progressing after docetaxel (TheraP ANZUP 1603). Journal of Clinical Oncology 40, 10–10 (2022).

20. S. Pathmanandavel et al., The prognostic value of post-treatment PSMA and FDG PET/CT in metastatic, castration-resistant prostate cancer treated with (177)LuPSMA-617 and NOX66 in a phase I/II trial (LuPIN). J Nucl Med, (2022).

21. O. Sartor et al., Lutetium-177-PSMA-617 for Metastatic Castration-Resistant Prostate Cancer. The New England journal of medicine 385, 1091–1103 (2021).

22. J. P. Buteau et al., PSMA and FDG-PET as predictive and prognostic biomarkers in patients given [^177^Lu]Lu-PSMA-617 versus cabazitaxel for metastatic castration-resistant prostate cancer (TheraP): a biomarker analysis from a randomised, open-label, phase 2 trial. Lancet Oncol 23, 1389–1397 (2022).

23. S. P. Thang et al., Poor Outcomes for Patients with Metastatic Castration-resistant Prostate Cancer with Low Prostate-specific Membrane Antigen (PSMA) Expression Deemed Ineligible for (177)Lu-labelled PSMA Radioligand Therapy. Eur Urol Oncol 2, 670–676 (2019).

24. S. Halabi et al., Prognostic model for predicting survival in men with hormone-refractory metastatic prostate cancer. J Clin Oncol 21, 1232–1237 (2003).

25. S. Halabi et al., Updated prognostic model for predicting overall survival in first-line chemotherapy for patients with metastatic castration-resistant prostate cancer. J Clin Oncol 32, 671–677 (2014).

26. O. Smaletz et al., Nomogram for overall survival of patients with progressive metastatic prostate cancer after castration. J Clin Oncol 20, 3972–3982 (2002).

27. F. Tang et al., Chromatin profiles classify castration-resistant prostate cancers suggesting therapeutic targets. Science 376, eabe1505 (2022).

28. J. M. Chan et al., Lineage plasticity in prostate cancer depends on JAK/STAT inflammatory signaling. Science 377, 1180–1191 (2022).

